# Inflammatory and immune markers in HIV-infected older adults on long-term antiretroviral therapy: persistent elevation of sCD14 and of proinflammatory effector memory T cells

**DOI:** 10.1101/2022.02.24.22271429

**Authors:** Makiko Watanabe, Mladen Jergovic, Lisa Davidson, Bonnie J. LaFleur, Yvonne Castaneda, Carmine Martinez, Megan J. Smithey, Raymond P. Stowe, Elias K. Haddad, Janko Nikolich-Žugich

## Abstract

HIV-positive patients whose viral loads are successfully controlled by active antiretroviral therapy (ART) show no clinical signs of AIDS. However, their lifespan is shorter compared to individuals with no HIV infection and they prematurely exhibit a multitude of chronic diseases typically associated with advanced age. It was hypothesized that immune system aging may correlate with, and provide useful biomarkers for, this premature loss of healthspan in HIV+ subjects. Here, we tested whether the immune correlates of aging, including cell numbers and phenotypes, inflammatory status and control of human cytomegalovirus (hCMV) in HIV-positive subjects on long-term successful ART (HIV+) may reveal increased “immunological age” compared to healthy, age-matched cohort (HC) in participants between 50 and 69 years of age. Specifically, we expected that younger HIV+ subjects may immunologically resemble older individuals without HIV. We found no evidence to support this hypothesis. While T cells from HIV+ participants displayed different expression of several differentiation and/or inhibitory/exhaustion markers in different T cell subpopulations, aging by a decade did not pronounce these changes. Similarly, while the HIV+ participants exhibited higher T cell responses and elevated inflammatory marker levels in plasma, indicative of chronic inflammation, this trait was not age-sensitive. We did find differences in immune control of hCMV, and, more importantly, a sustained elevation of sCD14 and of proinflammatory CD4 and CD8 T cell responses across age groups, pointing towards uncontrolled inflammation as a factor in reduced healthspan in successfully treated older HIV+ patients.

## INTRODUCTION

The development and widespread use of antiretroviral therapy (ART) has reduced AIDS-associated deaths dramatically since the late 1990s (Palella et al. 1998). Even though the life expectancy of HIV positive individuals increased, they exhibit higher rates of non-AIDS-related morbidity and mortality compared to HIV-negative counterparts (Guaraldi et al. 2011; Althoff et al. 2015; Rasmussen et al. 2015). Active HIV infection causes many abnormal immune phenotypes reminiscent of natural aging, including low CD4/CD8 ratio, chronic inflammation, immune activation and increase in immunosenescence markers (Sereti et al. 2017; Ronsholt et al. 2013; Kamat et al. 2012). While this analogy is tempting, it is also critically important to remember that HIV initially causes a catastrophic disruption of mucosal immunity and CD4 deficiency (Yamashita et al. 2001; Brenchley, Price, and Douek 2006; Appay et al. 2007), and while ART helps revert both of these manifestations, it is unclear whether full reconstitution of both parameters is ever achieved. Therefore, most of the HIV-related phenotypes correlate with virus load or decreased CD4 counts and poor disease prognosis in active HIV infection. Once the ART is initiated, as virus load decrease and CD4 count rises, abnormal immune phenotypes also get back to normal range (Gandhi et al. 2017). However, some of those immunological changes still exist in successfully treated HIV positive individuals including impaired memory T cell (van Grevenynghe et al. 2008) and B cell(van Grevenynghe et al. 2011) function, and loss of follicular helper T cell function(Cubas et al. 2015). Those phenotypes in HIV positives appear to be chronic inflammatory responses and they correlate with the functional status of the subjects, which includes parameters such as frailty and the immune risk factors (Castilho et al. 2016).

Immune phenotypes are sensitive to variations based on parameters that include, but are not limited to, age, gender, cytomegalovirus (CMV) serostatus. Human CMV (hCMV) is a widely distributed human herpesvirus that infects 60-90% of the human population depending on the age and socioeconomic conditions (Cannon, Schmid, and Hyde 2010; Korndewal et al. 2015; Zuhair et al. 2019). It is a latent persistent infection that is compatible with health in immunocompetent humans, but it represents a substantial threat to immunocompromised, as well as to the developing fetus(Emery 2001; Istas et al. 1995). It is known that many stressors, including third-party infection, will reactivate hCMV in a subclinical manner, causing initial viral gene transcription and translation, and, depending on the reactivating stimulus, also DNA replication and viremia (Cook et al. 2002; Forte et al. 2020; Prosch et al. 2000; Polic et al. 1998; Soderberg-Naucler et al. 2001). Human immune system devotes substantial resources to hCMV control, that can include up to 50% or more of circulating CD8 T effector memory (Tem) cells and T effector memory cells reexpressing CD45RA (TEMRA)(Munks et al. 2006; Sylwester et al. 2005). No other human microbial pathogen is known to modulate the immune system and host responses more extensively than hCMV – in a monozygotic twin study, it has been shown that 58% of the >100 immune parameters measured were modulated by hCMV infection (Brodin et al. 2015). Therefore, hCMV represents a bellwether measure of immune system fitness and is ideal to assess whether and how this virus is controlled under different physiological situations (Brodin et al. 2015; Rolle and Brodin 2016; Yan et al. 2021).

To assess the impact of controlled HIV infection and age on immune homeostasis and function, including hCMV control, we recruited HIV positive subjects from 50-69 years of age, who tested HIV-positive at least 5 years ago, and who were currently successfully treated with ART. To avoid sex at birth bias, we focused on male participants, and took advantage of the fact that nearly all HIV+ participants were CMV seropositive. We compared their immune and inflammatory phenotypes to those of HIV negative control groups with similar age, gender and CMV positivity. Consistent with prior studies, we found that the some of the measured parameters were significantly different in HIV+ subjects compared to HIV-negative controls. While many changes were not age-sensitive, we found age-dependent differences as well. Moreover, certain HIV-associated disturbances persisted in ART-treated participants into the advanced age, suggesting a prolonged impact of long-term chronic viral infection on proinflammatory immune status in older adults. These phenotypes may provide clues to overcome morbidity and mortality in successfully treated HIV-positive populations.

## RESULT

### Demographic characteristics of our cohort and study hypothesis

Our cohort consisted of 88 male participants aged 50-69, and were divided in groups of 17-25 participants according to HIV status and age, with 25 HIV+ and 24 HIV-participants aged 50-59 and 17+ and 22 HIV-participants aged 60-69 years, with very close medians between the comparing groups. All participants were verified to be serologically hCMV+. Demographic features of the cohort are shown in **Table 1**. The main hypothesis of our study was that HIV will be adding immunological “years” to the parameters studied, so that the younger HIV+ participants will resemble older HIV-participants in many immunological parameters.

**Table 1.** Demographics of study cohort

|  | Age 50-59 |  | Age 60-69 |  |
| --- | --- | --- | --- | --- |
|  | HIV+ (n=25) | HC (n=24) | HIV+ (n=17) | HC (n=22) |
| Age, years (range) | 55.4 (50-59) | 55 (50-59) | 64.4 (60-69) | 63.8 (60-69) |
| Years from diagnosis (range) | 20.4 (6-33) | N/A | 21.1 (6-35) | N/A |
| Ethnicity, n (%) |  |  |  |  |
| Hispanic or Latino | 9 (36%) | 9 (37.5%) | 3 (17.6%) | 12 (54.5%) |
| NOT Hispanic or Latino | 16 (64%) | 15 (62.5%) | 14 (82.4%) | 10 (45.5%) |
| Race, n (%) |  |  |  |  |
| Native American/Alaska Native | 0 (0%) | 1 (4.2%) | 0 (0%) | 1 (4.5%) |
| Asian | 0 (0%) | 3 (12.5%) | 0 (0%) | 0 (0%) |
| Black or African American | 3 (12%) | 2 (8.3%) | 3 (17.6%) | 0 (0%) |
| White | 19 (76%) | 16 (66.7%) | 14 (82.4%) | 20 (90.9%) |
| Unknown/Not Reported | 0 (0%) | 1 (4.2%) | 0 (0%) | 1 (4.5%) |
| Other | 2 (8%) | 0 (0%) | 0 (0%) | 0 (0%) |
| More than one | 1 (4%) | 1 (4.2%) | 0 (0%) | 0 (0%) |

### Systematic inflammatory markers in plasma

Persistent inflammation is one of the common phenotypes for HIV infection and aging and inflammatory markers in plasma are considered to be a good indication of the overall inflammatory status (Bektas et al. 2018; Ferrucci et al. 2002). We measured inflammatory markers in plasma from our cohort, including those reported to show consistent age-related increase in healthy HIV-uninfected aging cohorts, such as IL-6, CRP, soluble (s)TNF receptors 1 and 2, and TNFα; as well as those reported to be increased in HIV as potential markers of gut barrier damage and permeability, such as soluble CD163 and soluble CD14. IL-6, IL-8, sTNFR1, CRP (trend but no significance) and sCD163 levels were not significantly different between HIV-positive subjects (HIV+) and healthy controls (HC) in both age groups. TNF-α, sTNFR2 and sCD14 were significantly elevated in HIV+ compared to in HC (**Table 2**), yet behaved differently when analyzed across age groups. The difference between HIV+ and HC were significant only at age 50-59 for TNF-α (average and p value), only for age 60-69 for sTNFRII (average and p value), and in both groups for sCD14 (averages and p values). Of interest, in this cohort, none of the inflammatory markers were significantly higher in the older HC group, suggesting the elevation of those markers in HIV+ were likely related to HIV+ status and not to age-dependent inflammatory status.

**Table 2.** Inflammatory marker levels in plasma

|  | Age 50-59 |  |  | Age 60-69 |  |  |
| --- | --- | --- | --- | --- | --- | --- |
|  | HIV+ (n=25) | HC (n=24) | p value | HIV+ (n=16) | HC (n=22) | p value |
| IL-6 (pg/ml) | 7.19 ± 18.22 | 18.21 ± 32.56 | 0.155 | 1.33 ± 2.73 | 10.61 ± 39.31 | 0.354 |
| IL-8 (pg/ml) | 17.29 ± 24.06 | 22.02 ± 46.34 | 0.654 | 9.76 ± 6.53 | 20.32 ± 42.18 | 0.330 |
| TNF-α (pg/ml) | **27.91 ± 21.51** | **15.5 ± 10.53** | **0.014\*** | 14.94 ± 5.84 | 13.88 ± 4.62 | 0.536 |
| TNFR1 (ng/ml) | 1.06 ± 0.49 | 1.68 ± 1.95 | 0.143 | 0.93 ± 0.44 | 0.96 ± 0.56 | 0.878 |
| TNFR2 (ng/ml) | 5.93 ± 3.05 | 4.56 ± 2.46 | 0.092 | **5.87 ± 2.44** | **3.75 ± 2.14** | **0.008\*** |
| CRP (μg/ml) | 8.3 ± 11.67 | 11.41 ± 17.89 | 0.474 | 18.6 ± 27.02 | 5.14 ± 5.12 | 0.067 |
| sCD14 (μg/ml) | **2.7 ± 1.28** | **1.93 ± 1.11** | **0.031\*** | **2.57 ± 1.03** | **1.6 ± 0.87** | **0.003\*** |
| sCD163 (ng/ml) | 57.6 ± 36.52 | 60.16 ± 74.54 | 0.880 | 47.22 ± 35.26 | 39.15 ± 29.88 | 0.451 |

Data are mean ± standard deviation. Bold datasets and p value indicate p is less than 0.05. Abbreviations: IL, interleukin; TNF, tumor necrosis factor; TNFR, TNF receptor; CRP, C-reactive protein, sCD14, soluble CD14, sCD163, soluble CD163

### Reduction of CD4 T cells and increase of CD8 T cells in PBMCs from HIV+ subjects

To evaluate T cell homeostasis in HIV+ participants on ART as a function of aging, we first compared the absolute numbers of total T cells and their subsets in the peripheral blood mononuclear cells (PBMCs) from HIV+ and HC (gated as in **Fig. 1A**). Absolute number of CD4 T cells were lower and absolute number of CD8 T cells higher in HIV+ compared to HC, which resulted in the decrease of CD4/CD8 ratio (**Fig. 1B, C**). These changes were driven by likely HIV-mediated original depletion of CD4 Tn and Tcm subsets, and by an expansion of CD8 T cell subsets as a likely consequence of direct or bystander stimulation with HIV as well as by reactivating and/or opportunistic infections. The absolute number of γδ T cells was also significantly higher in HIV+ subjects **(Fig. 1D**). Of interest to our hypothesis, similar to the inflammatory marker changes, the changes in the numbers of CD4 and CD8 between HIV+ and HC were less significant in older group, probably due to the overall age-related lymphopenia. The fact that the aging did not potentiate the effects of HIV, but rather ameliorated the phenotype observed in HIV+, was surprising, and could suggest the frequency of cell phenotypes are affected differently by HIV infection and by age.

**Figure 1.**
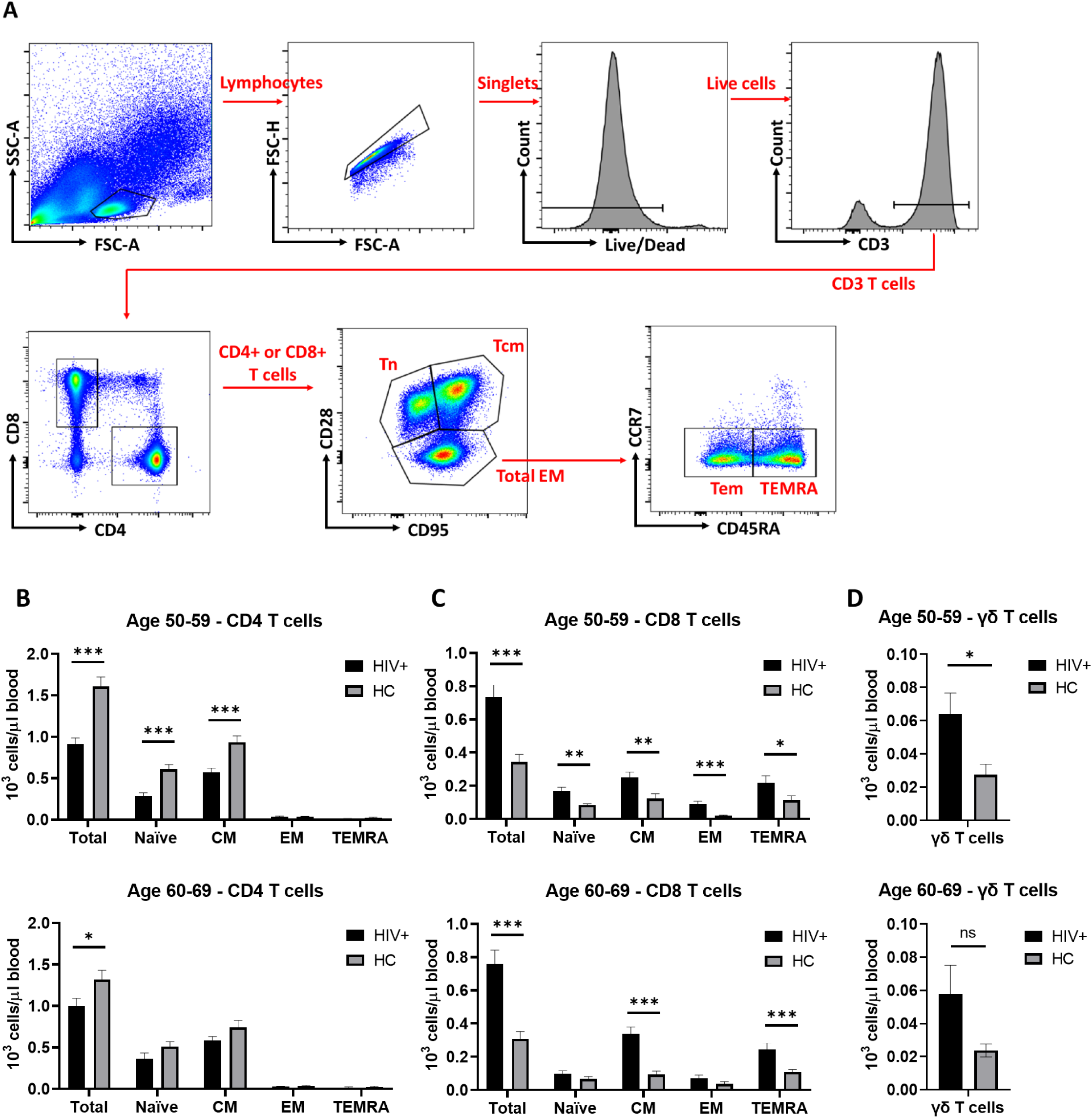
T cell subset analysis in PBMCs from successfully treated HIV positive patients (HIV+) and healthy control (HC). A) Gating strategy for CD4 and CD8 T cell subset analysis by flow cytometry shown in B. Lymphocyte population without cell debris was selected followed by removal of doublets and dead cells. Single positive CD4 and single positive CD8 cells are selected in CD3+ population and naïve, central memory (CM) and total effector memory (EM) cell populations are gated using CD28 and CD95, respectively. Total EM population was then divided into effector memory (EM) and CD45RA+ EM (TEMRA) using CCR7 and CD45RA. B) Absolute numbers of total, naïve, central memory (CM), effector memory (EM) CD4 and CD8 T cell subsets and δγ T cells in different age groups. Means and standard errors in each age/HIV group are shown. Significant differences between HIV+ (black) and HC (Grey) were determined by unpaired t-test. *p<0.05, **p<0.01, ***p<0.001.

### Expression of functional markers on T lymphocytes on successfully treated HIV positive subjects

To investigate the status of T lymphocytes in successfully treated HIV+, we measured expression of functional activation and inhibitory/exhaustion markers on CD4 and CD8 T cells. CD38 and HLA-DR have been recognized as activation markers expressed on activated cells during active microbial infection (Rodrigues et al. 2002). We found no difference in the frequencies of CD38+ or HLA-DR+ cells in CD4 or CD8 T cell subsets between HIV+ and HC participants of either age group (**Figs. 2A, B**). By contrast, the mean fluorescent intensity (MFI) of CD38 in CD38+ CD4 T cells was increased in HIV+ compared to HC in the 50-59 year olds. The difference became less significant because of the decrease of CD38 MFI over age (**Fig. 2G**). No significant differences we observed in CD38 MFI in CD38+ CD8 T cells, or HLA-DR MFI in either CD4 or CD8 T cells (**Figs. 2G, H**). We further evaluated expression of PD-1, TIM-3 and TIGIT as inhibitory markers, also expressed on exhausted T cells. We found no differences in the frequency of PD-1+ or TIGIT+ cells in CD4 or CD8 T cells between HIV+ and HC in our age groups (**Figs 2C, D**). However, PD-1 MFI increased in HIV+ in both age groups (**Fig. 2I**), whereas TIGIT MFI decreased in older HIV+ participants (**Fig. 2J**). By contrast, HIV+ younger (50-59 year old) T cells contained a significantly higher frequency of TIM3+ cells in both CD4 and CD8 T cells compared to age-matched HC counterparts (**Fig 2E**). In spite of the increase in TIM3+ cell frequency, its MFI in TIM3+ CD4 or CD8 cells decreased in HIV+ participants (**Fig. 2K**). Differences in TIM3+ cell frequencies or TIM3 MFI were not significant in older group (**Figs 2E, K**).

**Figure 2.**
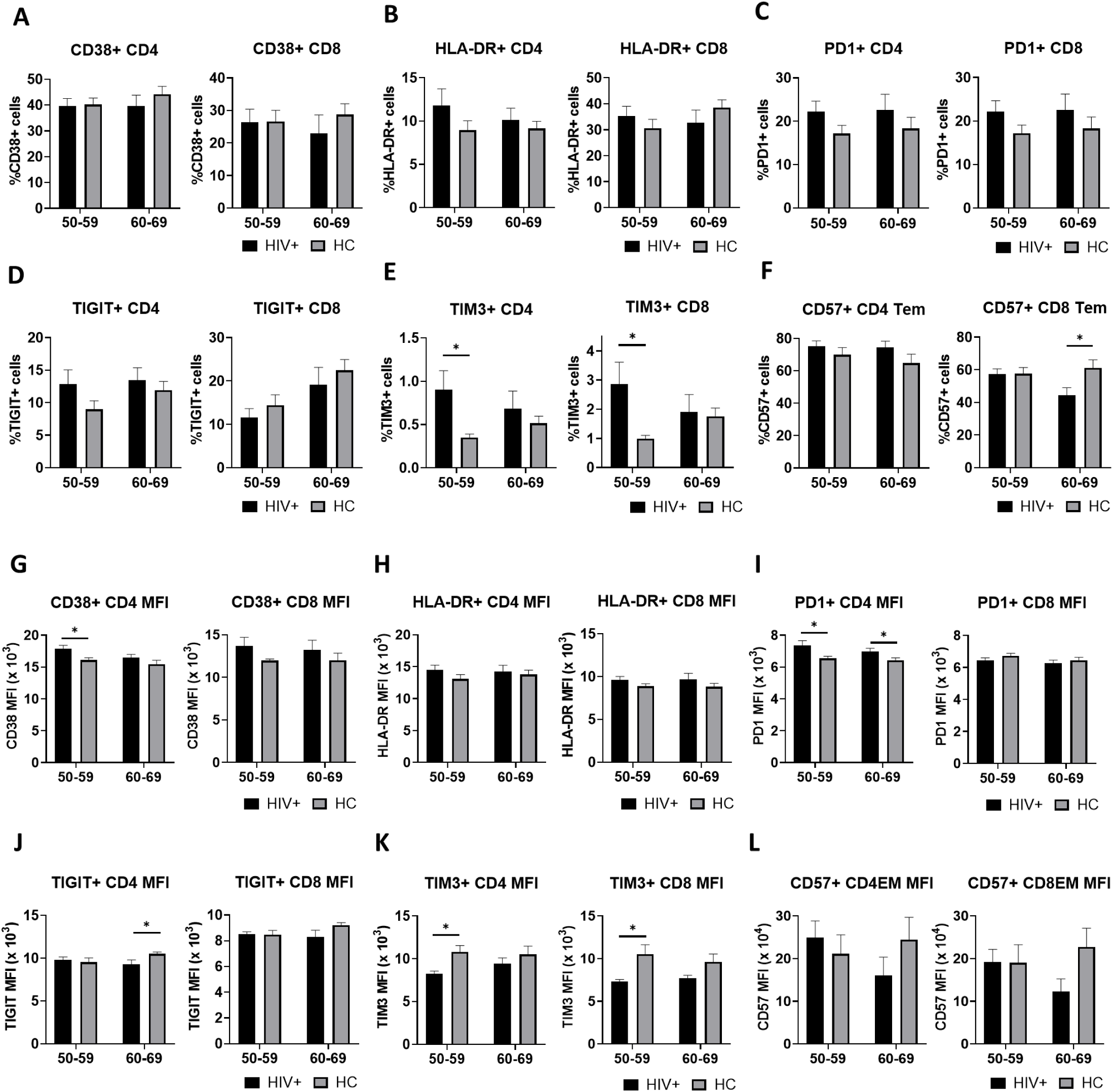
Expression of functional markers on CD4 and CD8 T cells in PBMCs from HIV+ and HC. A-F) frequency of CD38+ (A) and HLA-DR+ (B), PD-1+ (C), TIGIT+ (D), TIM3+ (E) and CD57+ (F) cells in total CD4 or CD8 T cells. G-K) Mean fluorescence intensity (MFI) of CD38 (G), HLA-DR (H), PD-1 (I), TIGIT (J), TIM3 (K), and CD57 (L) signals in each respective positive populations. Means (columns) and standard errors (bars) in each age/HIV group are shown. Significant differences between HIV+ (black) and HC (Grey) in each age were determined by unpaired t-test. *p<0.05.

CD57 is expressed on terminally differentiated cells, characterized by high levels of cytotoxicity and cytokine secretion and reduced or absent proliferation in response to TCR stimulation. The increase of CD57+ cells is often considered a correlate of T cell cellular senescence. In our dataset, we found that the older group of HIV+ subjects exhibited a decrease in frequencies of CD8 Tem cells (**Fig. 2F**). MFI analysis showed similar tendency and higher CD57 expression seems to be higher in CD8 Tem in the older group of HIV+ subjects, even though it is not statistically significant (p=0.057) (**Fig. 2L**).

Overall, the pattern of phenotypic changes suggested increased expression of inhibitory/exhaustion-related markers in HIV+ participants. Consistent with our other findings, such differences dominated in the younger age group, and the differences between HIV+ and HC diminished (or reversed some cases) amongst the older age group.

### T cell proliferation and functional markers on non-proliferative and proliferative cells

To evaluate functional characteristics of T cell subsets in successfully treated HIV+ population as a function of aging, we stimulated PBMC with the CD3/CD28/CD49d antibody cocktail to analyze T cell proliferative ability. Approximately 50% of CD4 T cells and 60% of CD8 T cells underwent at least one cell division. The number of proliferating cells did not differ between HIV+ and HC or between age groups in either CD4 and CD8 T cells, indicating that T cell in successfully treated HIV+ participants retain proliferative capacity at least under our experimental conditions (**Fig. 3B**). Analysis of expression of activation/inhibitory markers on cells at different number of cell divisions revealed that both PD-1+ cells and TIM3+ cells were less frequent amongst CD4 and CD8 cells that divided two or more times specifically in HIV+ participants and these data were significant only in the older age groups (**Figs 3C, D**). Careful data examination suggested, however, that cells from HIV+ participants exhibited relatively high levels of PD-1 in the first division, but that these levels were not sustained in more advanced division, unlike in their HC counterparts (**Fig. 3C**). Whether and to what extent that may belie different signaling via the TCR in HIV+ vs HC participants remains to be examined. Induction of TIM-3 was more gradual in both HIV+ and HC groups, being again similar in the first division, but then remaining constant on cells from HIV+ participants, and further increasing to much higher levels in the HC group (**Fig. 3D**), suggesting different regulation of these two markers and their different dysregulation in HIV+ carriers. The expression of TIGIT did not follow this pattern and exhibited somewhat increased levels in non-dividing and early-dividing CD4 T cells of HIV+ younger age group, but here HC cells caught up with TIGIT expression in the more advanced division (**Fig. 3E**). This difference was not observed in CD8 T cells, nor in the older age group.

**Figure 3.**
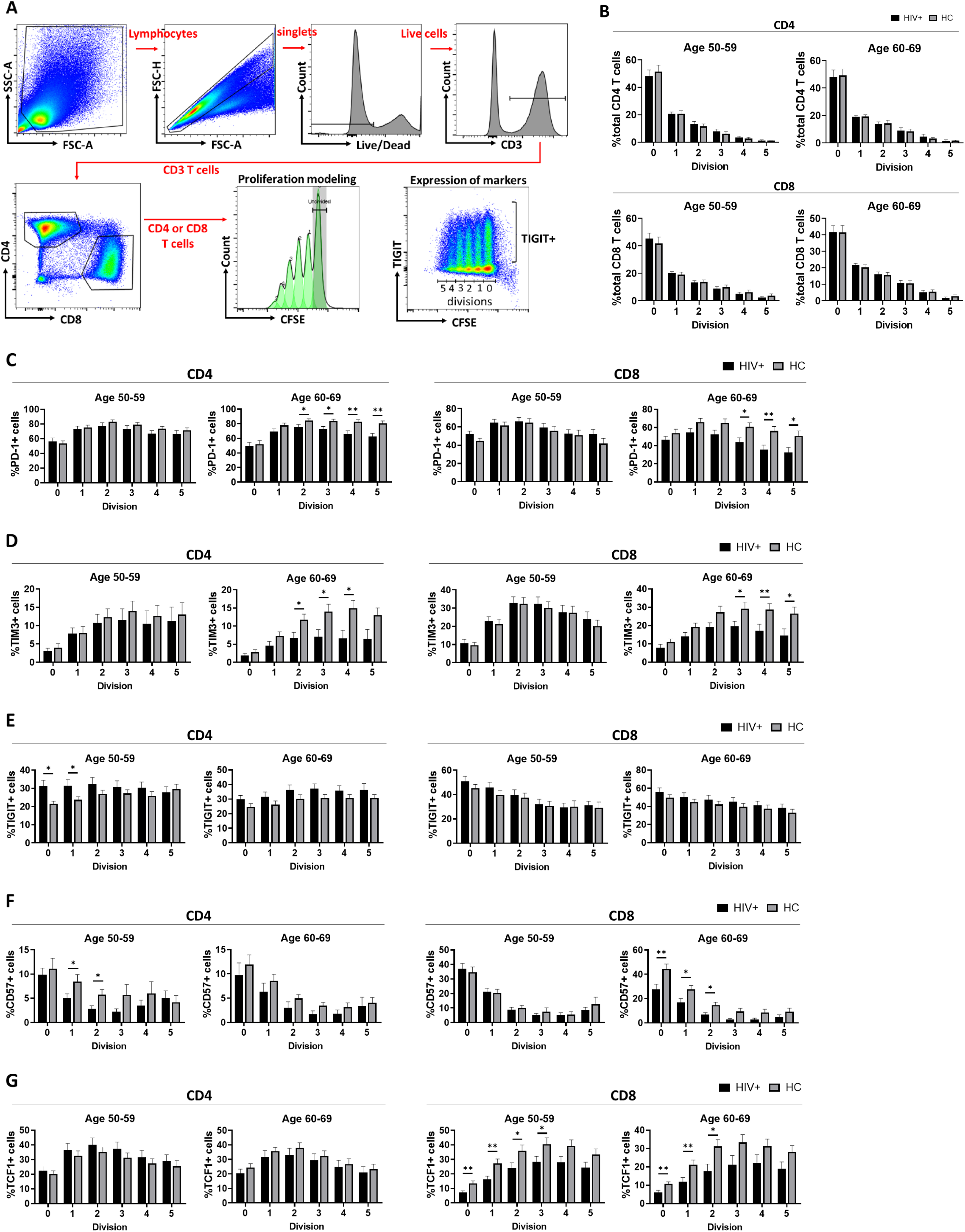
T cell proliferation and marker expression profiles in non-dividing and dividing CD4 and CD8 T cells after CD28/CD3/CD49d stimulation in HIV+ and HC. A) Gating strategy for non-dividing and dividing CD4 and CD8 T cells using proliferation modeling with FlowJo. The low-right panel shows gating example of dividing cells and positive populations on TIGIT. B) number of cells in each division (div). C-G) frequency of PD-1+ (C), TIM3+ (D), TIGIT+ (E), CD57+ (F) and TCF-1+ (G) cells in each divisions. Means (columns) and standard errors (bars) in each age/HIV group are shown. Significant differences between HIV+ (black) and HC (Grey) were determined by unpaired t-test. *p<0.05, **p<0.01.

Not surprisingly, the expression of CD57+ declined in proliferating T cells, and the kinetics of the decline was often faster in the HIV+ group compared to HC; significance of these changes was not uniform, and was present in CD4 cells from our younger and CD8 cells from our older group, usually in early divisions (**Fig. 3F**). TCF-1 is a self-renewing marker and its expression positively correlates with the replicative ability of T cells (Kratchmarov, Magun, and Reiner 2018). The TCF-1+ CD8 T cells were less abundant in HIV+ compared to HC groups in both age categories, consistent with the idea that the CD8 T cell compartment was stimulated more intensely in the HIV+ participants (**Fig. 3G**). Those result demonstrated that even though the bulk proliferative ability upon the stimulation did not appear to be altered by ART-controlled HIV infection, the proliferating cells exhibited increased expression of late differentiation markers. This suggests that T cells are altered during aging in the successfully treated HIV+ participants in numerous and subtle ways relative to HC counterparts.

### HIV+ individuals have higher humoral and cellular immune response against CMV

We took advantage of the fact that all our participants were hCMV-seropositive to examine how immune responses to CMV may be affected by HIV and aging. The hCMV-specific total antibody titers in plasma were significantly higher in HIV+ compared to HC younger participants (**Fig. 4A**). This difference was not significant in the older age group, likely due to the increase of CMV antibody titers in HC, whereas the HIV+ group did not exhibit such an increase (**Fig. 4A**). To examine functional capabilities of anti-hCMV humoral responses, we devised a hCMV fluorescence-based neutralization assay and determined the neutralizing antibody titers as described in the materials and methods. This analysis showed that hCMV neutralizing antibody titers remained constant and generally between low and negative in the HC group, even though all of the subjects had substantial overall antibody titers (**Fig. 4B**). HIV+ participants had significantly higher nAb titers than their HC counterparts, and the difference appeared to further increase with age. Furthermore, the correlation analysis showed that the total antibody titer and nAb titer did not correlate in HC but correlated significantly in HIV+ (**Figs. 4C, D**).

**Figure 4.**
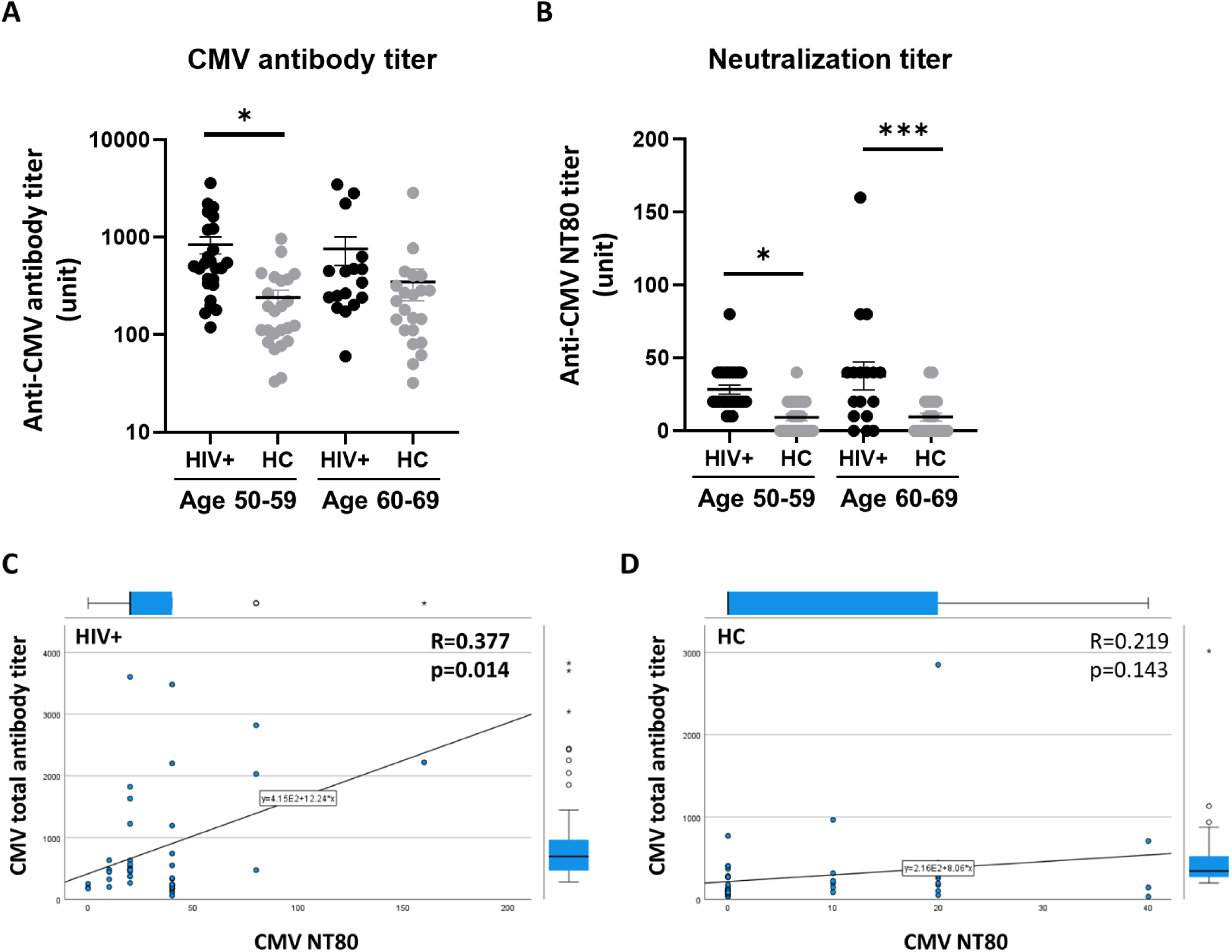
Anti-CMV antibody titers in HIV+ and HC plasma. A-B) Total antibody titers (A) and 80% Neutralization titers (B) against CMV in plasma. Significant differences between HIV+ (black) and HC (Grey) antibody titers were determined by unpaired t-test. *p<0.05, ***p<0.001. C-D) Correlation between total anti-CMV antibody titer and anti-CMV neutralization titer in HIV+ (C) and in HC (D). Pearson’s R and p values are shown in the graphs.

Given that the primary control of reactivating hCMV is under purview of T cells, we examined T cell cytokine secretion after both hCMV-specific (IE1 and pp65 peptide pools) and polyclonal, non-specific stimulation with phorbol ester and calcium ionophore (PMA and ionomycin). We measured IFN-γ and TNF-α expression by intracellular cytokine staining and scored the cells expressing either one or both cytokines as responsive. No significant difference was observed in % T cell reactivity to IE-1 in either CD4 or CD8 T cells from HIV+ and HC (**Fig. 5A, B**). By contrast, pp65-stimulated CD8 T cells were significantly more frequent in HIV+ participants than in HC in the younger age group (**Fig. 5A, B**). This difference was not present in the older group, where the HC CD8 T cells were as frequent as those in HIV+ participants (**Fig. 5 A, B**). We interpret these differences to suggest different levels of hCMV reactivation/replication at different ages, as discussed below

**Figure 5.**
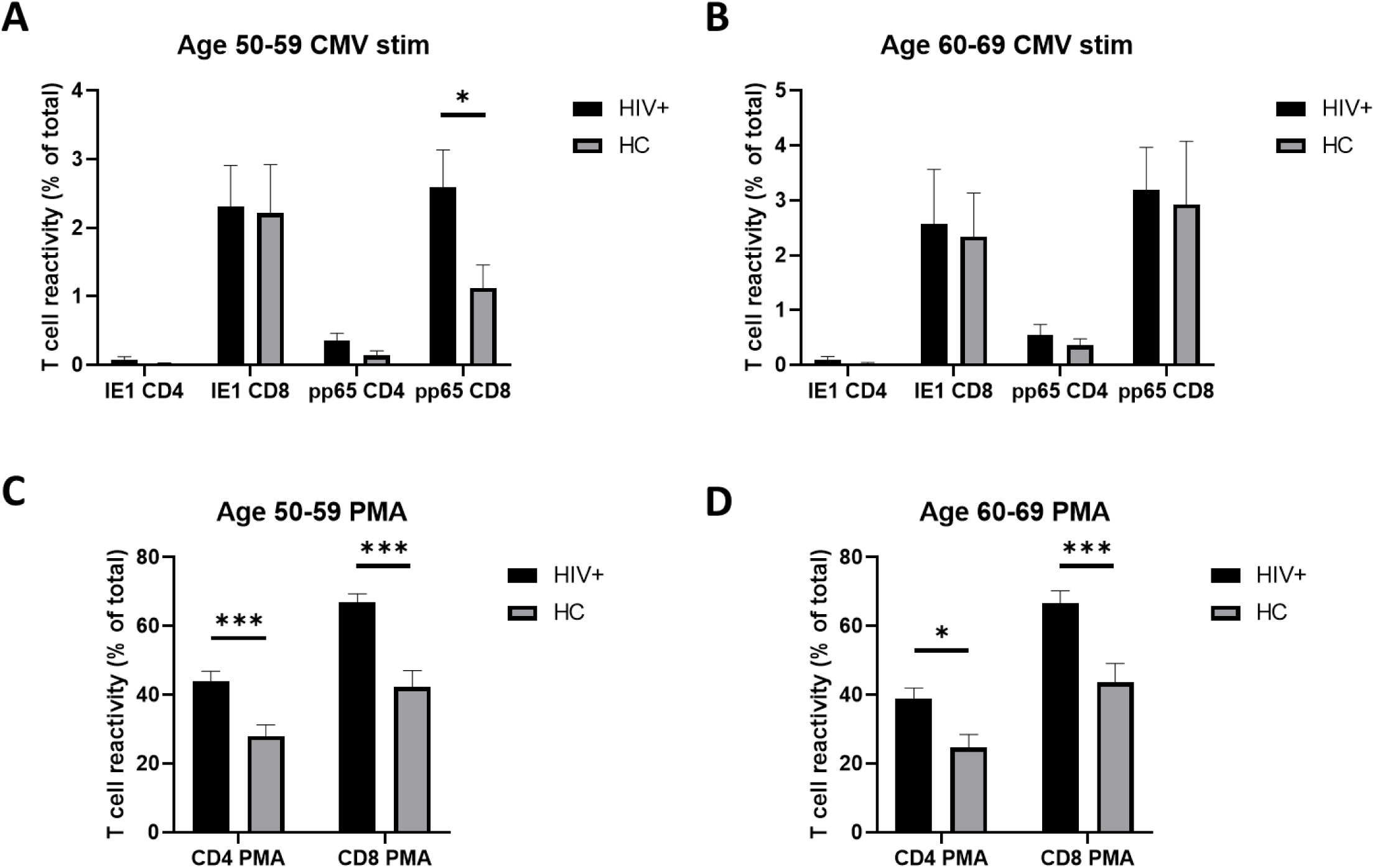
Cytokine response in T cells after in vivo stimulation. A-B) Frequency of IFN-γ+ and/or TNF-α+ cells after 3 hours of CMV-peptide stimulation of PBMCs from HIV+ and HC at age of 50-59 (A) or age of 60-69 (B). C-D Frequency of IFN-γ+ and/or TNF-α+ cells after 3 hours of PMA/ionomycin stimulation of PBMCs from HIV+ and HC at age of 50-59 (A) or age of 60-69 (B).

The response of T cells to PMA/ionomycin revealed higher cytokine responses in HIV+ than HC, a result that was consistent in both CD4 and CD8, and in both age groups (**Fig. 5C, D**). This broad increase in cytokine-secreting T cells may contribute to the pro-inflammatory status in HIV+ participants but does not appear to be age-sensitive at least in our participant cohort.

### HIV+ display distinct correlation pattern between inflammatory signatures and immune phenotypes different from HC

Finally, we analyzed the correlation between variables which displayed difference in HIV+ compared to HC to get the landscape of immune status in HIV+. **Figure 6** shows the correlation between the variables in HIV+ (**Fig. 6A**) and HC (**Fig. 6B**) participants. The certain categories (plasma inflammatory markers or marker expression on CD4 and CD8) correlated well with one another regardless of HIV status. Negative correlation of the cytokine+ T cells after PMA stimulation and TCF-1+ CD8 T cells was also commonly observed in both HIV+ and HC. Other correlation patterns greatly differed between HIV+ and HC. Thus, in HC, the inflammatory cytokines in plasma significantly correlated with the frequency of CD57+ T cells and cytokine+ T cells after the stimulation. Those correlations were generally absent in HIV+ participants, and instead, we found correlations between inflammatory cytokines and the frequency of TIM3+ T cells in HIV+, which was absent in HC.

**Figure 6.**
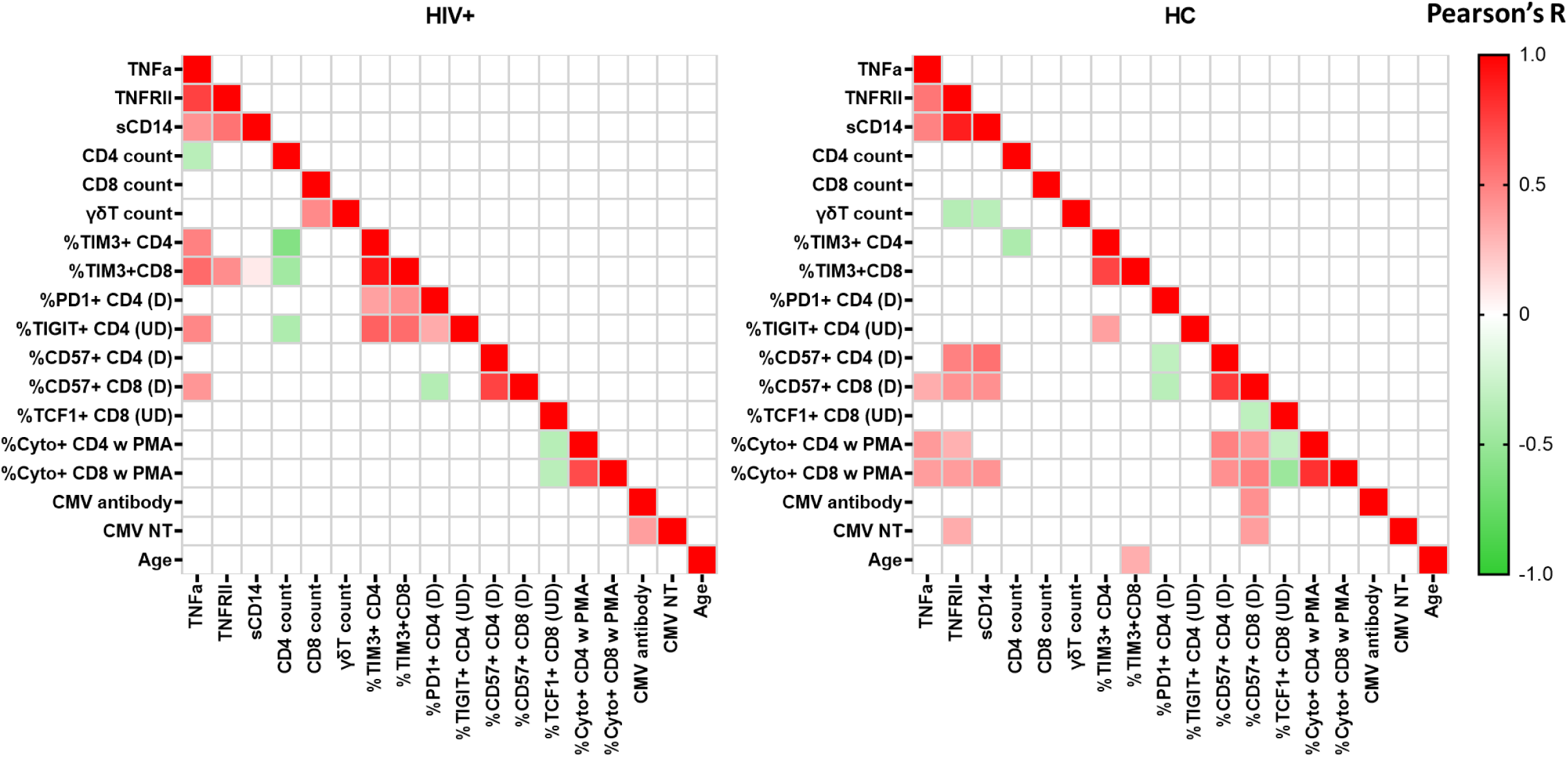
Heatmap of the bivariate Pearson correlation matrix. Correlation values are represented as colors. Red and green are the perfect linear positive and negative relationships, respectively. White means p-value is less than 0.05. Left: HIV+, Right: HC. (D): divided cells, (UD): undivided cells

## DISCUSSION

HIV infection causes a massive and catastrophic disruption in the host immune system and mucosal homeostasis, which in subjects on ART resemble some of the immune changes found in physiological aging. It remains difficult to prove whether those homeostatic disturbances in HIV+ participants actually represent accelerated aging or whether they only result in phenotypes similar to aging. Active HIV infection causes many changes in immune phenotypes such as CD4 T cell death, T cell activation and exhaustion and expression of inflammatory markers. Those changes are significantly reduced with ART, yet, chronic diseases abound in this group of patients and appear earlier than in their HIV-negative counterparts. Some of diseases were attributed to ART toxicity especially in the early days of ART (Group et al. 2007; Ryom et al. 2013; Scherzer et al. 2012), but the ART has been improved and the increase of those diseases over time does not exclude low HIV replication as a cause. The chronic diseases in HIV-positive patients with successful ART correlates with inflammatory markers, while those markers do not correlate with duration of ART (Burdo et al. 2011; Kuller et al. 2008; Sandler et al. 2011). Therefore, the high mortality and morbidity by non-AIDS related disease in HIV-positive patients with ART is more likely due to HIV infection itself, rather than ART toxicity.

In this study, we performed immune profiling of successfully treated (at least 5 years of ART and <50 copies HIV RNA per ml plasma) HIV+ participants and age matched HIV-negative controls, all seropositive for hCMV. Our focus in this study was to understand how virologically undetectable HIV infection affects immune aging under successful ART. We were somewhat surprised to find that systemic inflammatory markers in those individuals were subtly and not very predictably elevated, and in particular that IL-6 and CRP, the two stalwart markers of elevated inflammation with aging, did not change in an age-related manner over the span of two decades in our cohort. However, our data were similar to the findings of de Armas et al., who compared plasma inflammatory cytokines from successfully treated HIV+ with those from HC in different age groups and found only TNFR2 was elevated in HIV+ in all age groups while IL-6 and IL-8 were actually decreased in HIV+ compared to HC (de Armas et al. 2017). Moreover, Margolick et al., reported that TNF-α was the only analyte out of 17 measured cytokines or chemokines (including IL-6, IL-8 and CRP) which showed significant increase in successfully treated HIV+ group compared to HC participants (Margolick et al. 2018). In our study, the only sustained inflammation-related marker elevated in both younger and older HIV+ participants relative to their HC counterparts was sCD14, generally viewed to be a surrogate marker for microbial translocation. The persistent elevation of sCD14 after years of ART treatment (Mendez-Lagares et al. 2013) and association of sCD14 plasma level and all-cause mortality in HIV+ with ART (Sandler et al. 2011) suggests the sCD14 may be a biomarker of chronic diseases in HIV+ with ART, and, perhaps, that impaired gut barrier function may be one of the factors driving these conditions..

Major T cell subsets were predictably reduced (CD4 Tn and Tcm) or elevated (all CD8 subsets) with successfully treated HIV infection, and these changes showed no no age-sensitivity. T cell phenotypes in successfully treated HIV positive subjects were not very different from those in HC, lacking signs of massive activation or massive cellular exhaustion, and only showing increased expression of Tim-3 in the 50-59 year old groups. However, even though the T cell proliferation in response to polyclonal stimulation was not globally altered by HIV status, the signs of altered T cell function were found in reduced expression of PD-1 and TIM-3 on cells undergoing second and further division. TIGIT was expressed on many more non-dividing CD4 T cells in HIV+ compared to HC participants, and frequency of TCF-1+ (self-renewing) cells was lower in CD8 T cells of HIV+ compared to HC at early divisions. However, even these changes were not age-sensitive, and many of them were either unchanged or attenuated in the older groups.

Immune response to hCMV was more informative and insightful and suggested impaired control of hCMV in HIV+ participants, with possible increased rate of virus reactivation based on the fact that HIV+, but not HC, participants exhibited significant increase in anti-hCMV neutralizing antibodies, and that this increase was more pronounced in the older groups. T cell responses to hCMV immediate-early antigen 1 (IE1) regardless of the HIV status and ages. As its name states, IE1 is amongst the earlies transcripts of hCMV and is transcribed early both in the course of primary infection and reactivation (Ye et al. 2020), explaining ubiquitous and strong T cell stimulation by this antigen. T cell responses against pp65, a late-transcribing major tegument phosphoprotein (Ye et al. 2020), were more pronounced in the HIV+ younger group, consistent with the idea that the higher cytokine response in HIV+ participants likely reflects strong hCMV reactivation/ precipitated by the HIV infection itself. The finding that older HC older subjects “catch up” with HIV+ participants in their T cell anti-pp65 responses is likely reflective of the repeated rounds of hCMV subclinical reactivation over time in both groups. Finally, analysis of cytokine production following polyclonal stimulation revealed strong, persistent propensity of T cells from HIV+ participants to produce significantly more inflammatory cytokines (IFNγ and TNFα) across age groups.

Multiparametric correlation analysis between all of the analyzed traits has allowed us to evaluate distinct correlation profiles between inflammatory markers, T cell counts and frequencies, phenotypes and functional responses in successfully treated HIV+ and HC participants. The inflammatory markers elevated in HIV+ did not correlate with T cells responses, but rather positively correlated with frequency of inhibitory factor/exhaustion markers (TIGIT and TIM3) on CD8 cells, and negatively correlated with CD4 counts, consistent with damage to CD4 cells, exhaustion of CD8 cells and incompletely repaired gut barrier. This pattern of correlation was dramatically different in HC, where inflammatory cytokines positively correlated with T cell responses to polyclonal stimulation and to expression of CD57, suggesting qualitative difference in immune system regulation.

Limitations of this study are primarily twofold. First, the study is of relatively low power, with 17-25 subjects/group, which could distort findings in both experimental and control groups. Second, the age-related conclusions and the results on age-sensitive traits are limited by a relatively narrow span of ages, from 50-69 years of age, split into decade-based bins for comparison purposes. The fact that over this range of ages we did not observe changes in certain well-known age-sensitive parameters, like IL-6 and CRP levels, may highlight that type of limitation. However, even with these limitations, we have identified two sustained and mechanistically potentially different contributors to increased systemic inflammation in HIV+ participants, with a potential to exacerbate multiple chronic diseases. These include: (i) the persistent significantly higher elevation of sCD14; and (ii) increased frequencies of T cells producing inflammatory cytokines (IFNγ and TNFα) upon polyclonal stimulation. Both were elevated across both ages examined, suggesting prolonged disturbances in gut mucosal integrity and potential microbial translocation, and a linked, or independent/additive, chronic “trigger-happy” inflammatory T cell accumulation, both of which could contribute to the adverse effects of poorly controlled inflammation and its impact on various chronic diseases (Bektas et al. 2018; Sandler and Douek 2012).

## EXPERIMENTAL PROCEDURES

### Human subjects and sample preparation

This study was approved by the Institutional Review Boards (IRB) at the University of Arizona (IRB protocol#2102460536). HIV positive subjects, who are treated successfully with ART (HIV RNA <50 copies/ml in plasma), were recruited from Banner-University Medicine North HIV positive study cohort was selected from HIV-positive males, who diagnosed more than 5 years ago, and are over 50 years old of age. Age and sex-matched controls are healthy community-dwelling individuals, recruited in Arizona. CMV seronegative individuals were removed from the cohort since CMV infection affects many aspects of immune phenotype. Demographic information of the cohort is shown in Table 1. Blood for complete blood count was collected in BD vacutainer with EDTA and submitted to Sonora Quest. Blood for peripheral blood mononuclear cells (PBMCs) and plasma was collected in BD Vacutainer with sodium heparin. Plasma was separated by centrifugation at 1100 x g for 10 minutes and PBMC was isolated from the buffy coat by Ficoll-Paque PLUS (GE Healthcare) and cryopreserved in fetal bovine serum (FBS) + 10% DMSO.

### Measurement of inflammatory markers

Interleukin-6 (IL-6), Interleukin-8 (IL-8) and tumor necrosis factor (TNF)-α (HCYTOMAG-60K and HSTCMAG-28SK), TNF receptor (TNFR) 1 and 2 (HSCRMAG-32K), soluble CD14 (sCD14) (HCVD6MAG-67K), and C-reactive protein (CRP) (HNDG2MAG-36K) in plasma were quantified by Millipore MagPix kit (Millipore) and soluble CD163 in plasma was quantified by ELISA kit (Invitrogen EHCD163) according to the manufacturer’s protocol.

### Antibody Titers against CMV

Serum samples with high IFA-scored antibody titers (i.e., 2560), obtained from prior studies, were used as the top standards for CMV as previously described (Stowe 2014). Two-fold serial dilutions of the standards (2560, 1280, 640, 320, 160, 80, 40, and 20) were made with PBS in separate tubes. One hundred microliters of positive and negative controls, standards, and diluted patient samples (all dilutions were at 1:101 with PBS) were pipetted in duplicate into individual microplate wells followed by a 30 min incubation (all steps were carried out at room temperature). The plates were then washed 3 times with 350 μl wash buffer using an Embla microplate washer (Molecular Devices, Menlo Park, CA). Next, 100 μl of enzyme conjugate (peroxidase labeled anti-human IgG) was pipetted into the wells followed by another 30 min incubation period. The plates were then washed 3 times, and 100 μl of chromogen substrate (TMB/H2O2) was pipetted into the wells. The plates were then covered to protect from direct light and incubated for 15 min. One hundred microliters of 0.5 M sulfuric acid was added to each well to stop the reaction. Absorbance was then read at 450nm (reference wavelength 620nm) using a SpectraMax Plus 384 (Molecular Devices). The values of the unknown samples were assigned in relation to the standard curve.

### Neutralization antibody titer to CMV

Human foreskin fibroblast (HFF)-1 was purchased from ATCC (#SCRC-1041). HFF-1 cells were grown in complete DMEM (4.5g glucose/L, 4mM L-glutamine, 1mM Sodium Pyruvate, 1X Penicillin/Streptmycin and 15% FBS). GFP-expressing hCMV virus was kindly provided by Dr. Felicia Goodrum (University of Arizona)(Umashankar et al. 2011). Human plasma from each subject was diluted in complete medium at 1:10, followed by two-fold dilutions 5 times. The diluted plasma was then mixed with same volume of medium containing 800 infectious unit (IU) of virus and incubated for 2 hours at room temperature. HFF-1 monolayer in a 96-well plate was infected with half amount of the plasma/virus mixture. No plasma control wells were infected with medium containing 400 IU of the virus. The plate was incubated in 37 °C CO2 incubator for 48 hours. The number of GFP positive cells per field was counted by Cytation 5 (Biotek). Cut-off value was calculated as 20% of no plasma control, which represent 80% reduction of the infection. The highest dilution rate, which exceeded 20% positive, was read as a neutralizing titer.

### PBMC Stimulation and Flow Cytometry

Cryopreserved PBMCs were thawed in RPMI medium supplemented with 10% FBS, penicillin and streptomycin in the presence of DNAse (Sigma, Saint Louis, MO), rested overnight in X-Vivo medium (Lonza/Basel, Switzerland) supplemented with 5% human male AB serum. Antibodies used for flow cytometry is listed in supplemental table 1. For proliferation experiment, PBMCs were stained with CFSE Cell Proliferation kit (Invitrogen) and cultured in a plate coated with antibodies against CD3 (BioLegend), CD28 (eBioscience) and CD49d (BD Pharmingen), in the presence of IL-2 (TECIN™ Teceleukin, National Cancer Institute) at 100 unit/ml for 5 days. Fresh medium was added every 2 days. Following incubation cells were stained with surface staining antibodies for 1h in PBS (Lonza) + 2% FCS at 4 ^0^C. Dead cells were stained with Live/Dead Fixable Blue Dead Cell Stain Kit (Invitrogen) and cells were fixed and permeabilized with FOXP3/Transcription Factor Staining Buffer Set (Invitrogen) for intracellular staining (ICS) of TCF-1. For CMV peptide or PMA/Ionomycin stimulation, PBMCs were stimulated with CMV pp65 and IE1 PepTivators (Miltenyi Biotec) or Cell Stimulation Cocktail (Invitrogen) in the presence of Brefeldin A (Millipore Sigma) for 4 hours. Dead cells were stained with Zombie Aqua (BioLegend) and all cells were fixed and permeabilized with BD Cytofix/Cytoperm (BD) for ICS of cytokines.

The subsets of T cells are defined as followed; Naïve (Tn): CD28^int^CD95^low^; central memory (Tcm): CD28^high^CD95^high^; effector memory (Tem): CD28^low^CD95^high^CCR7^-^CD45RA^-^ terminally differentiated EM (Te); CD28^low^CD95^high^CCR7^-^CD45RA^+^ (Figure 1A). The gate for expression of functional markers were set by using a fluorescence minus one controls. Samples were acquired using a Cytek Aurora cytometer (Cytek) and analyzed by FlowJo™ v10.7.2 Software (BD Life Sciences).

### Statistics

Unpaired t-test and simple bivariate correlation analysis was performed using IBM SPSS Statistics for Windows, version 28 (IBM Corp., Armonk, N.Y., USA)

## Data Availability

All data produced in the present study are available upon reasonable request to the authors

## ACKNOWLEDGEMENTS

We thank the healthcare providers at Banner-University Medical Center for their assistance in the recruitment of the participants, and Dr. David Harris and staffs at Arizona Health Sciences Center Biorepository for the biospecimen processing and biobanking. We also thank Dr. Felicia D. Goodrum and her lab members for providing TB40/E-5 clone for hCMV neutralizing assay.

## AUTHOR CONTRIBUTIONS

M.W., M.J., L.D., M.J.S. and R.P.S. conducted experiments and acquired data. M.J., M.J.S. and J.N.Z. contributed to study design. M.W. and B.J.L analyzed and interpreted the data. M.W. Y.C. and C.M. performed recruitment, specimen collection and database maintenance. M.W., E.K.H. and J.N.Z. contributed in writing the manuscript.

## DATA AVAILABILITY STATEMENT

The datasets generated during and/or analyzed during the current study are available from the corresponding author on reasonable request.

## PATIENT CONSENT STATEMENT

All patients provided written informed consent to the study

